# Changes in Frequency of Resuscitation Among the Oldest Old Following Japan’s End-of-Life Care Guideline Revision: A Population-Level Interrupted Time-Series Analysis Using National Open Claims Data

**DOI:** 10.64898/2026.05.28.26354307

**Authors:** Michi Sakai, Takeo Nakayama

**Author notes:** Corresponding author: Michi Sakai, Yoshida konoe-chou Sakyo-ku, Kyoto city, Kyoto, Japan.

## Abstract

Resuscitation in the oldest old at the end of life is associated with potential harm, raising concerns about misalignment with patients’ goals of care. This study aimed to elucidate changes in the use of resuscitation among the oldest old in Japan following the revision of the national guideline on end-of-life care which explicitly incorporates the concept of advance care planning. We conducted a repeated cross-sectional study using the National Database of Health Insurance Claims Open Data, including adults aged ≥85 years, from April 2014 to March 2024. The annual number of resuscitation procedures per 100,000 individuals aged ≥85 years was used as the measure of frequency. Resuscitation included closed-chest cardiopulmonary resuscitation (CPR) and endotracheal intubation. Interrupted time series analysis was used to examine changes following the 2018 revision of the national end-of-life care guideline. The frequencies of CPR and endotracheal intubation declined before 2018 (CPR: age 85-89, −68.4 [−87.9 to −48.8]; age ≥90, −106.7 [−131.5 to −82.0]; intubation: age 85–89, −57.5 [−71.8 to −43.2]; age ≥90, −69.5 [−80.7 to −58.3]), but the decline attenuated thereafter (CPR: age 85–89, +56.2 [28.0 to 84.5]; age ≥90, +84.1 [50.7 to 117.6]; intubation: age 85–89, +36.6 [8.5 to 64.7]; age ≥90, +38.3 [23.8 to 52.8]). These findings provide insight into the changes in resuscitation trends following policy interventions supporting end-of-life decision-making. Further studies are needed to better understand the mechanisms underlying this change.

## Introduction

Japan is the world’s earliest example of a super-aged, high-mortality society, with adults aged 85 years and older comprising 4.8% of the population and annual deaths exceeding 1.6 million in 2024 (1). The intensity of care at the end of life among the oldest old, those aged 85 years or older, has become a major social concern, as higher-intensity care may impose substantial burden while compromising the quality of end-of-life care (2–4). In particular, resuscitation in the oldest old at the end of life is associated with potential harms (5) and may be misaligned with patients’ goals of care (6–13). Survival to hospital discharge is approximately 12 −15% after in-hospital cardiac arrest (6) and approximately 3% after out-of-hospital cardiac arrest in patients aged ≥80 years (7–9). As health status deteriorates and death approaches, older patients tend to forgo CPR when provided with appropriate information and structured communication (10–12). Recent national surveys indicate a decline in the proportion of the general public who would opt for CPR at the end-of-life (13). These findings highlight the importance of structured support for decision-making about resuscitation among the oldest old (14). The 2025 American Heart Association guidelines state that resuscitation decisions should not be based solely on age and emphasize the importance of establishing patient-centered goals of care (15). In Japan, the Ministry of Health, Labour and Welfare issued the Guidelines for the Decision-Making Process of End-of-Life Care in 2007 and revised them in March 2018 to explicitly incorporate the concept of advance care planning (ACP). In the same year, ACP was incentivized through reimbursement for end-of-life care, thereby promoting its implementation in clinical practice (16).

Evidence suggests that policy interventions designed to enhance end-of-life decision support are associated with changes in resuscitation preferences among the oldest old (17–19). Following the introduction of physician reimbursement for advance care planning under the Affordable Care Act in 2016, the use of do-not-resuscitate (DNR) orders increased significantly among patients aged 85 years or older with heart failure in the United States (19). After the mandate for Physician Orders for Life-Sustaining Treatment (POLST) registration was introduced in Oregon, registrations increased among persons aged 95 years and older (17). In addition, the introduction of the Veterans Affairs Life-Sustaining Treatment Decisions Initiative in 2018 - which seeks to standardize the decision-making process for patients with serious illness and poor prognoses - was accompanied by increased documentation of treatment preferences among those aged 90 years and older (18). In contrast, in South Korea, following the implementation of the Life-Sustaining Treatment Decisions Act in 2018 - which legally permits the withholding or withdrawal of treatment including CPR - individuals aged 80 years and older demonstrated lower levels of self-determination, and documentation of their preferences remained limited (20).

Nevertheless, the magnitude of nationwide changes in resuscitation use among the oldest old following policy interventions to support end-of-life decision- making remains poorly understood. Observational studies have shown a recent decline in end-of-life resuscitation among older populations (21, 22). Our previous study in Japan reported a decreasing trend in the use of CPR and mechanical ventilation among the oldest old in the period after the initial publication of the Guidelines for the Decision-Making Process of End-of-Life Care in 2007 (23); however, changes after the 2018 revision of the guideline have not been evaluated. This study aimed to elucidate changes at the national level in the frequency of resuscitation among the oldest old following the 2018 revision of the Guidelines for the Decision-Making Process of End-of-Life Care in Japan, to provide insights into its potential impact.

## Materials and Methods

### Study design and subjects

We performed a repeated cross-sectional study to elucidate changes in the frequency of resuscitation among individuals aged 85 years and older following the March 2018 revision of Japan’s Guidelines for the End-of-Life Decision-Making Process.

### Data source and measures

We utilized the National Database of Health Insurance Claims Open Data (NDB-OD) (24) and vital statistics (25), both released by the Ministry of Health, Labour and Welfare of Japan. The NDB-OD is a publicly available dataset derived from the National Database of Health Insurance Claims and Specific Health Checkups of Japan (NDB) (26). It provides annual aggregated counts of each procedure performed, stratified by sex and 5-year age categories. The NDB contains nearly all digitized health insurance claims, thereby enabling nationwide examination of real-world medical practices that can be readily assessed. Data from the 2014 fiscal year (FY) to the 2023 FY were available for analysis as of October 30, 2025. Vital statistics were used to obtain the population counts for each age group and each fiscal year.

The annual number of resuscitation procedures performed per 100,000 individuals aged ≥85 years was used as the measure of frequency. We examined closed-chest-CPR and endotracheal intubation as resuscitation. Each procedure was identified using procedure codes (J-codes) utilized for the Japanese national fee schedule. To minimize potential double counting when procedures were performed multiple times in a single individual, we used the code for the initial day of closed-chest CPR (J046) and the code for endotracheal intubation used for resuscitation purposes (J044).

### Data analysis

Interrupted time series analysis using segmented regression was conducted to estimate the effect of the guideline revision on resuscitation use among the oldest old. This approach enables rigorous quasi-experimental evaluation of population-level health interventions (27). The interruption time (i.e., the time of policy intervention, corresponding to the guideline revision) was set at March 2018 (i.e., FY 2017). To control for time-varying confounding, changes in end-of-life care utilization during the COVID-19 pandemic (28) were included in the regression model. Because the data were available annually, the number of time points was limited; therefore, a level-change term was not included to avoid overfitting given the limited degrees of freedom.

Regression models of the following form were developed:

Y_t = β_0 + β_1·time_t + β_2·time_after_t + γ·covid_post_t + ε_t

Where

Y_t = the number of resuscitation procedures performed per 100,000 individuals aged ≥85 years at each fiscal-year time point;

time_t = FY_t - 2014 (the number of years since the start of the study);

time_after_t = max (0, FY_t - 2018) (the number of years after the intervention);

covid_post_t = indicator variable that equals 1 for fiscal years 2020 and later to denote the COVID-19 period, and 0 otherwise;

*ε*_t = error term;

The coefficients assessed to determine the effect of guideline revision are:

*β*_0 = intercept;

*β*_1 = slope in the pre-intervention period;

*β*_2 = change in slope after the intervention (i.e., the difference between pre-and post-intervention slopes);

*γ* = coefficient representing the level shift associated with the COVID-19 pandemic;

Newey-West heteroskedasticity- and autocorrelation-consistent (HAC) standard errors were used to account for serial correlation and heteroskedasticity in the residuals.

### Sensitivity analysis

A controlled interrupted time series (CITS) analysis (29) was performed using comparison procedures to account for concurrent secular trends. CITS incorporates a comparison series that is not exposed to the intervention, enabling adjustment for system-wide time-varying factors and isolation of the intervention-specific effect. Cataract surgery was used as the comparison series because it is widely performed among the oldest old, is minimally affected by seasonality or long-term fluctuations, and is unlikely to be directly influenced by the revision of end-of-life care guidelines while sharing external factors unrelated to the intervention. We modeled the log ratio of resuscitation to cataract surgery frequencies per 100,000 as the outcome, defined as: R_t = log(Resuscitation_freq_t / Cataract_freq_t).

Where

Resuscitation_freq_t = the number of resuscitation procedures performed per 100,000 individuals aged ≥85 years at each fiscal-year time point;

Cataract_freq_t= the number of cataract surgery procedures performed per 100,000 individuals aged ≥85 years at each fiscal-year time point;

The CITS model was specified as

R_t = β_0 + β_1·time_t + β_2·time_after_t + γ·covid_post_t + ε_t

The estimated coefficients were converted to relative percent changes using exp(β)-1. This approach is algebraically equivalent to modelling the difference in trends between the intervention and control series (29).

We additionally performed a sensitivity analysis in which the interruption time (i.e., the time of guideline revision) was defined as FY 2018 to account for uncertainty in assigning the timing of the guideline revision within fiscal year–based data. R version 4.5.2 (2025-10-31) was used for the analysis. We set the significance level of each test to 5%.

### Artificial intelligence disclosure

Artificial intelligence (AI) tools (e.g., ChatGPT, OpenAI; GPT-4 and GPT-5 models) were used to assist with literature search and organization, generation of statistical analysis code in R, and improvement of the manuscript. All outputs were critically reviewed and verified by the authors, and all decisions regarding study design, analysis, and interpretation were made solely by the authors, who take full responsibility for the content.

## Results

The annual changes in the number of resuscitation procedures performed per 100,000 individuals aged ≥85 years from FY 2014 to 2023 are presented in Fig 1 and Table 1. Comprehensive details on the number of procedures performed per 100,000 individuals aged ≥85 years in each fiscal year are provided in the Supporting information 1. The frequencies of CPR and endotracheal intubation declined before 2018, but this decline attenuated thereafter (CPR: age 85–89, +56.2 [28.0 to 84.5]; age ≥90, +84.1 [50.7 to 117.6]; intubation: age 85–89, +36.6 [8.5 to 64.7]; age ≥90, +38.3 [23.8 to 52.8]) (Table 1). The attenuation of decline was larger in magnitude among individuals aged ≥90 years.

**Figure 1.**
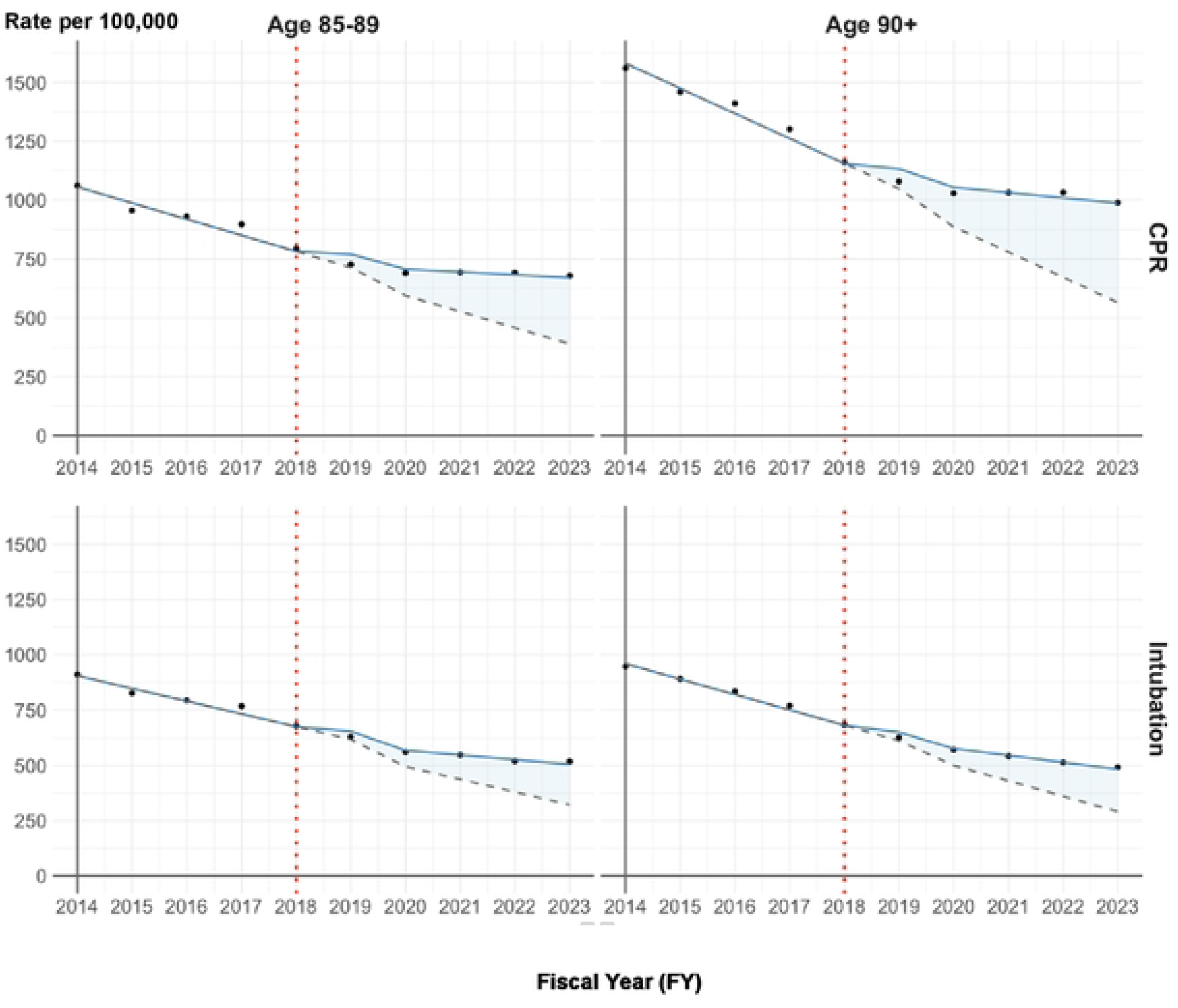
Change in the number of resuscitation procedure cases per 100,000 population.

**Table 1.**
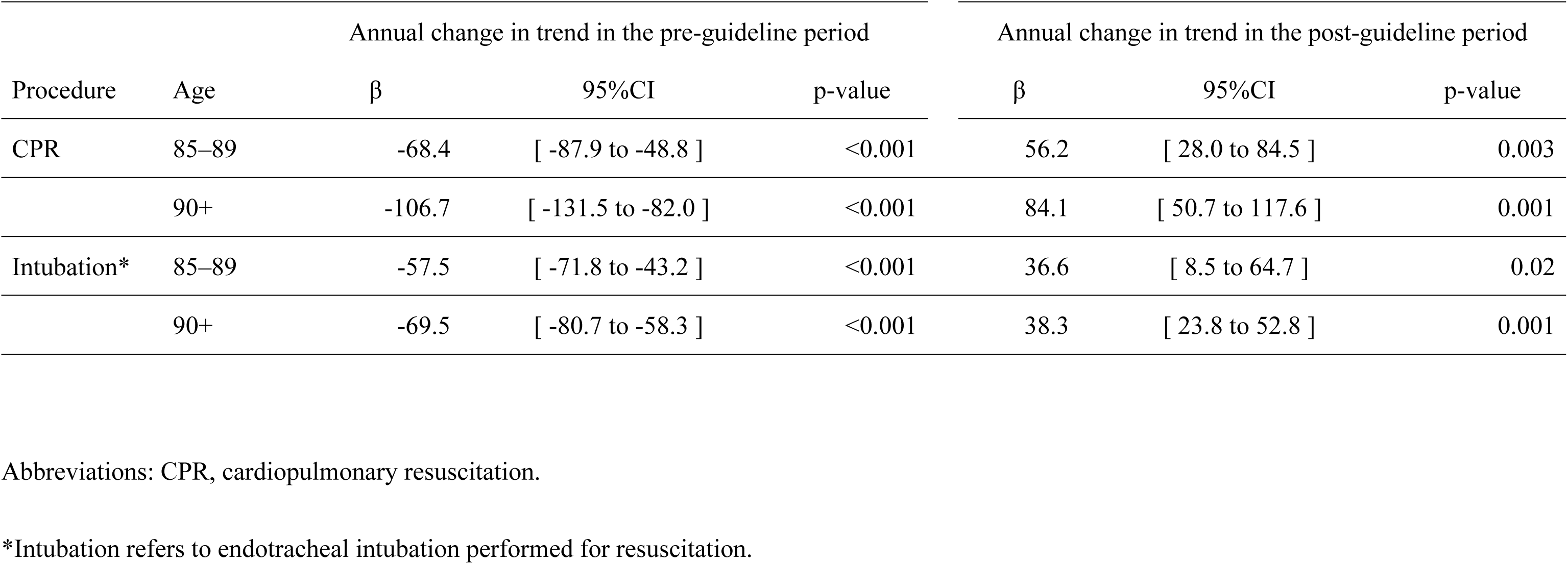
Changes in the number of resuscitation procedure cases per 100,000 population among the oldest-old following Japan’s 2018 national end-of-life guideline revision.

| Procedure | Age | Annual change in trend in the pre-guideline period |  |  | Annual change in trend in the post-guideline period |  |  |
| --- | --- | --- | --- | --- | --- | --- | --- |
| | | $\beta$ | 95%CI | p-value | $\beta$ | 95%CI | p-value |
| CPR | 85–89 | -68.4 | [ -87.9 to -48.8 ] | <0.001 | 56.2 | [ 28.0 to 84.5 ] | 0.003 |
|  | 90+ | -106.7 | [ -131.5 to -82.0 ] | <0.001 | 84.1 | [ 50.7 to 117.6 ] | 0.001 |
| Intubation* | 85–89 | -57.5 | [ -71.8 to -43.2 ] | <0.001 | 36.6 | [ 8.5 to 64.7 ] | 0.02 |
|  | 90+ | -69.5 | [ -80.7 to -58.3 ] | <0.001 | 38.3 | [ 23.8 to 52.8 ] | 0.001 |

Abbreviations: CPR, cardiopulmonary resuscitation.
\*Intubation refers to endotracheal intubation performed for resuscitation.

The results of the controlled interrupted time series are shown in Table 2. The extent of the reductions in CPR relative to the control procedure was attenuated in the post-intervention period, and no statistically significant decreases were detected (age 85-89, −1.2% [−4.7% to 2.4%]; age ≥90, −2.2% [−7.1% to 2.9%]). In contrast, intubation showed a statistically significant relative decrease, although the upper bound of the confidence interval approached 0% (age 85-89, −3.5% [−6.7% to −0.2%]; age ≥90, −5.0% [−9.4% to −0.5%]). The results were also generally consistent in the analysis defining the interruption time as FY2018, with the direction and magnitude of post-intervention slope changes remaining consistent across specifications.

**Table 2.**
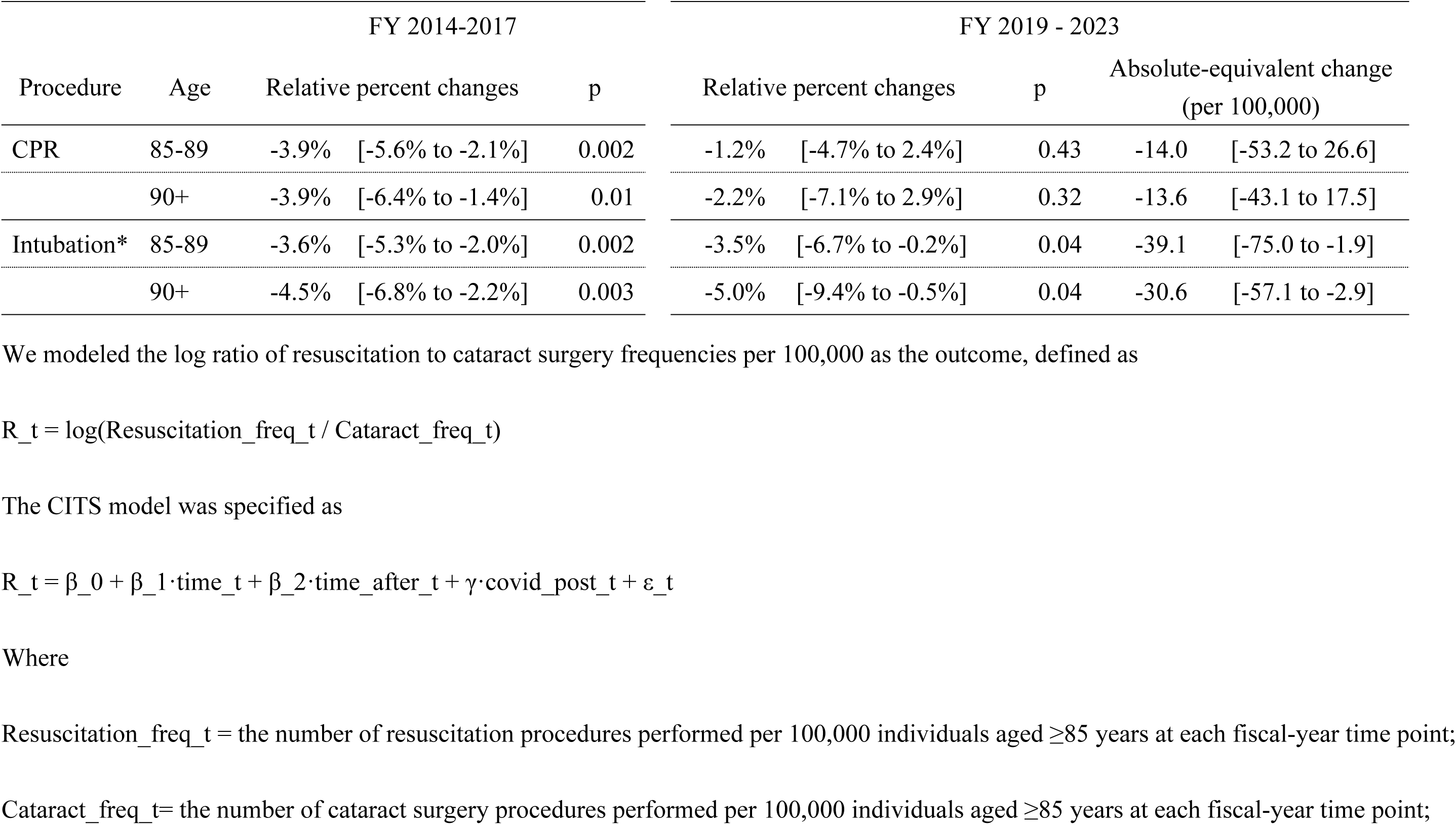
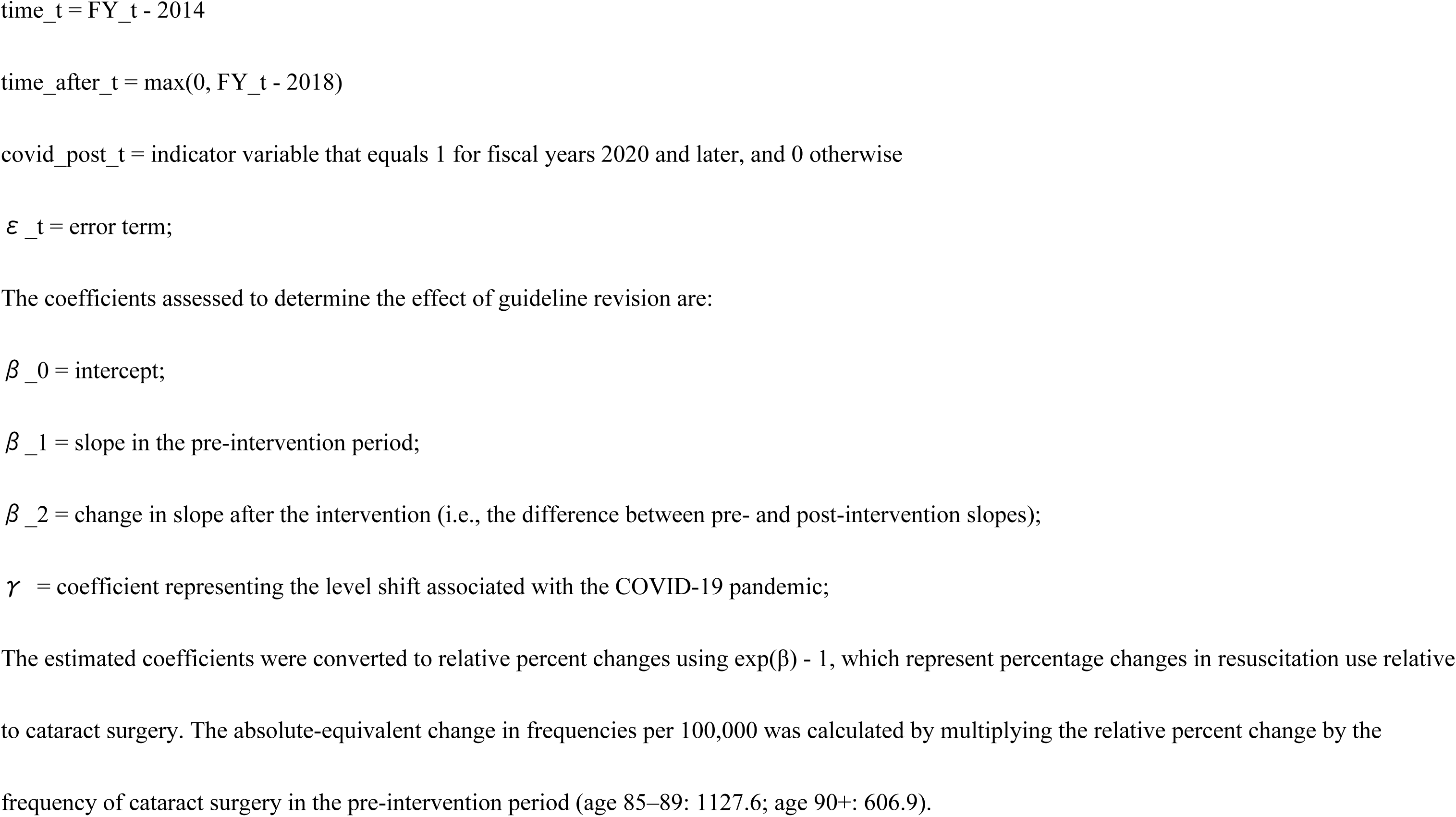
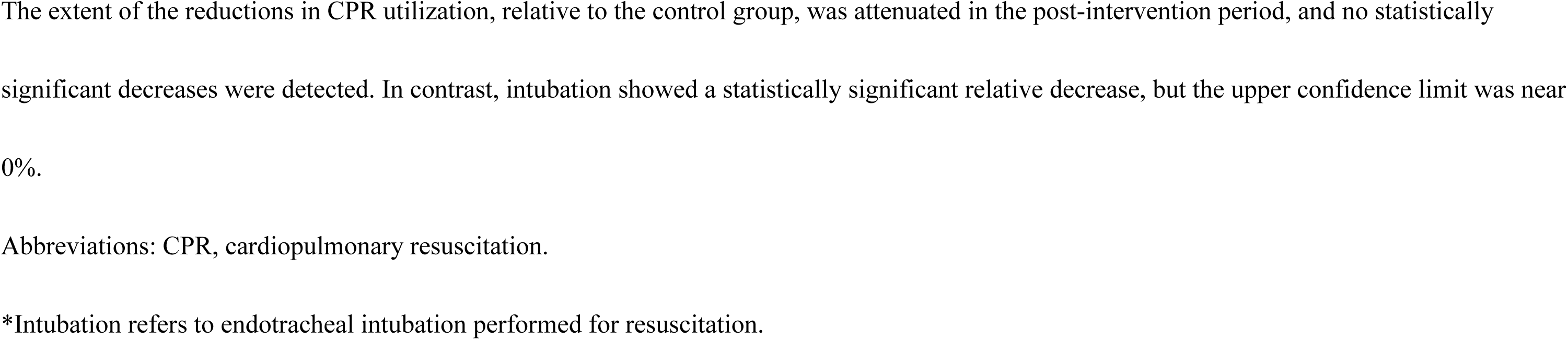
Sensitivity analysis using cataract surgery as a control.

## Discussion

Following the revision of the guidelines in March 2018, the decline in the frequency of CPR and endotracheal intubation attenuated. To our knowledge, this is the first study to examine nationwide changes in resuscitation among the oldest old following policy interventions for end-of-life decision making. These findings provide insights into changes in temporal trends in resuscitation practices following the guideline revision.

There are several possible explanations for why the decline attenuated after the guideline revision. First, resuscitation had already been declining before the 2018 revision of the guideline. We previously reported a decrease between 2012 and 2014 (23), suggesting that the decline had begun following the initial issuance of the guideline in 2007, and that the incremental impact of the 2018 revision may therefore have been limited. This tendency is consistent with prior research evaluating institutional interventions aimed at supporting decision-making for life- sustaining treatment (30, 31). No decline in CPR use among adult inpatients was observed in South Korea after the implementation of the Life-Sustaining Treatment Decisions Act (30), possibly because CPR performed against patients’ wishes was already uncommon before the Act (32). In the United Kingdom, the introduction of the Recommended Summary Plan for Emergency Care and Treatment (ReSPECT) was not followed by a reduction in futile CPR, likely because such CPR had already been declining beforehand (31). Second, it is possible that engagement in end-of-life care planning did not increase sufficiently, and thus subsequent changes in clinical practice did not emerge. In Japan, public recognition of ACP has remained low even after the guideline revision (33). This is consistent with findings from prior studies examining the effects of policies supporting end-of-life decision-making. Uptake of ReSPECT process remained low following its introduction in the United Kingdom (31), and completion of POLST forms remained limited after implementation of the Life-Sustaining Treatment Decisions Act in South Korea (30). There is a potential gap between the dissemination of ACP and changes in clinical practice (34). Further research is needed to examine changes in the decision-making process. Finally, because the NDB-OD is available only as annual aggregated values with a limited number of observation points, it is possible that the limited number of observations in the time series reduced the statistical power to detect changes associated with the intervention. The Cochrane Effective Practice and Organization of Care (EPOC) group recommends that interrupted time-series studies include at least three data points both before and after the intervention in order to be eligible for inclusion in a review (35). In this study, the pre-intervention period comprised four annual data points (fiscal years 2014-2017), and the post-intervention period was limited to five data points, thereby satisfying the EPOC criteria for interrupted time-series designs.

Among those aged ≥90 years, the decline during the period before the guideline revision was greater than in those aged 85–89 years for both closed-chest CPR and intubation. The attenuation of the decline after the guideline revision was also greater, possibly reflecting the magnitude of the earlier decrease. An alternative explanation is that the attenuation of the post-intervention decline may reflect a floor effect, whereby further reductions in the use of resuscitation procedures become increasingly limited as their frequency declines to lower levels. Prior studies suggest that older populations are less engaged in decision-making, and that interventions to support decision-making may have a more limited impact on treatment preferences in these groups (20, 36, 37). Our findings provide limited evidence of age-related differences in changes in temporal trends during the post-intervention period. Differences in the underlying trend by age group were nevertheless observed. Further investigation is warranted to examine age-related differences in the impact of guideline revisions.

Several limitations of this study should be noted. Given the limitations of the NDB Open Data and the absence of a clearly defined level change, our findings should be interpreted as reflecting changes in temporal trends rather than causal effects of the guideline revision. The NDB Open Data provides annual counts of treatment procedures performed; individual patient data are not available. Consequently, while frequencies of resuscitation use in the total population can be measured, frequencies among end-of-life patients cannot be identified. These frequencies should be interpreted as a proxy measure rather than an indicator of end-of-life care intensity, limiting our ability to evaluate the impact of the guideline revision on resuscitation practice. Furthermore, because the data were available only on an annual (FY) basis, the number of time points was limited. This may have reduced statistical power and led to an underestimation of true changes. Moreover, abrupt level changes immediately after the policy intervention, as well as other temporal fluctuations, may not have been captured. The denominator of the frequency of resuscitation use was defined as the annual number of procedures performed. Although a single individual could, in principle, undergo multiple procedures within a year, potentially leading to overestimation, this approach may overestimate the frequency of use. However, the present study focused on procedures that are unlikely to be performed repeatedly for the same individual, and therefore any potential overestimation is expected to have minimal impact on the assessment of temporal trends. In addition, because individual patient data were not available, we could not ensure that the control series was subject to the same co-interventions or external events as the intervention series. Although cataract surgery was selected as a control series due to its high frequency and relative stability, it may still have been influenced by healthcare system factors, such as capacity constraints, particularly during the COVID-19 period. Therefore, the validity of the control series assumption cannot be fully guaranteed. Finally, our results are applicable in Japan, and generalizability is limited.

Despite these limitations, the availability of granular data on end-of-life care among the oldest old remains limited and represents a growing global challenge (38, 39). In recent years, open government health data have become increasingly available across countries, with growing expectations for the democratization of medical research and the facilitation of policy impact assessment using readily available data (40–43). Open data allow for continuous monitoring and identification of signals of change and key research gaps, complementing studies based on patient-level data.

In conclusion, the decline in resuscitation among the oldest old attenuated following the 2018 revision of the guideline. These findings provide insight into changes in resuscitation trends following policy interventions supporting end-of-life decision-making. Further studies are needed to better understand the mechanisms underlying this change.

## Type of contribution of the authors

Conceptualization: MS, TN; Investigation: MS, Methodology: MS, TN; Software: MS; Formal analysis: MS, TN; Writing—original draft preparation: MS, TN; Supervision: TN; Resource: TN;

All authors have read and agreed to the published version of the manuscript.

## Acknowledgment

Not applicable.

## Data availability

Data were derived from publicly available sources: the National Database of Health Insurance Claims Open Data (NDB-OD) and Vital Statistics, both provided by the Ministry of Health, Labour and Welfare of Japan.

The NDB-OD is available at: https://www.mhlw.go.jp/stf/seisakunitsuite/bunya/0000177182.html

Vital Statistics are available at: https://www.mhlw.go.jp/toukei/list/81-1a.html

## Ethical consideration

Ethical review was not required since the NDB-OD did not contain any identifiable personal information.

## Patient consent statement

Not applicable.

## Clinical trial registration

Not applicable.

## Supporting information

**S1 Table.** The number of resuscitation procedure performed per 100,000 population among the oldest-old (2014–2023)

